# Sexual risk-taking behavior amongst emerging adults in a tertiary institution of learning in Coastal Kenya: A qualitative study of stakeholders’ perspectives using causal loop mapping

**DOI:** 10.1101/2023.04.06.23288135

**Authors:** Stevenson K. Chea, Vincent A. Kagonya, Eunice A. Oyugi, Carophine Nasambu, Isaac Menza, Fauz Ibrahim, Osman Abdullahi, Alice Anika, Amin S. Hassan, Souheila Abbeddou, Kristien Michielsen, Amina Abubakar

## Abstract

**Background:** It is known from previous studies that university students in sub-Saharan Africa (sSA) engage in sexual risk-taking behaviour (SRTB). However, there is paucity of data on correlates of SRTB among university students (emerging adults {EmA}) at the Kenyan Coast thus hindering intervention planning. This study seeks to provide an in-depth qualitative understanding of correlates of SRTB and their interconnectedness among university students at the Kenyan coast combining qualitative research with a systems thinking approach.

**Methods:** Using the ecological model, and employing in-depth interviews, we explored the perceptions of twenty-one EmA and five other stakeholders on what constitutes and influences SRTB among EmA at a tertiary institution of learning in Coastal Kenya. Data were analysed using a thematic framework approach. A causal loop diagram (CLD) was developed to map the interconnectedness of the correlates of SRTB.

**Results:** Our findings show that unprotected sex, transactional sex, cross-generational sex, multiple sex partnerships, gender-based violence, sex under influence of alcohol/drugs, early sex debut, and sharing sex toys were common SRTBs. Based on the ecological model and CLD, most of the reported risk factors were interconnected and operated at the individual level followed by those that operate at social level.

**Conclusion:** Our study shows that EmA are mostly engaging in unprotected sex. Enhancing sexuality education programs for students in Kenyan universities and strengthening support systems including counselling for those using alcohol/drugs may help reduce SRTB among EmA in universities in Kenya.

## Background

Sexual risk-taking behaviour (SRTB) including non-condom use [1–4], concurrent sexual partners [5, 6], multiple sexual partnerships [1, 5, 7–11], early sex debut [5, 10–12], age-disparate relationships [3, 7, 9] and transactional sex [1, 3, 6, 11, 13] is well documented among young people in sub-Saharan Africa (sSA) and remains common [14]. Accordingly, sSA bears the greatest burden of HIV infection among young people [15], with young women and adolescent girls accounting for one in four new infections in 2019, despite making up only 10% of the total population [15]. Although progress has been made towards scaling down the HIV pandemic, Kenya remains one of the high burden countries in sSA [16]. In 2019, a total of 41,408 people were newly infected with HIV in Kenya, with 15-29 year old contributing 62% of all new infections [16].

Emerging adulthood is a new conception of development for the period from the late teens through the twenties, with a focus on ages 18-25 [17]. Emerging adults’ (EmA) brains are still developing [18], which increases risk for sub-optimal performance on executive function with a heightened propensity for engaging in SRTB as a consequence [18]. Further, concerns of sex tourism and drug abuse reported at the Kenyan Coast put EmA in this region at a higher risk of SRTB [19, 20]. According to the ecological model, the mechanisms that underlie SRTB involve multiple explanatory domains [21]. The model proposes that risk and protective factors of SRTB among young people fall into six domains: 1) macro, ii) social, iii) school, iv) family, v) peers, and vi) individual [21]. Additionally, there has been less attention to the causal mechanisms underpinning the occurrence of SRTB among EmA. Systems thinking enables understanding of inter-relationships and interactions within a system [22]. Causal loop diagrams (CLDs) are a systems thinking method that can be used to unpack complex health system behaviour [23]. The current study utilizes CLD to map the system surrounding SRTB.

Exploring the experiences of EmA and opinions of other stakeholders on SRTB is important in designing targeted interventions. Previous studies show that EmA in tertiary institutions of learning in sSA have a higher propensity for non-condom use [10, 24–42], multiple sexual partnerships [10, 24–38, 43], early sex debut [10, 26, 28, 30, 32, 36, 38, 44], transactional sex [10, 28, 30, 32, 37], age-disparate relationships [26, 32, 36] and concurrent sexual relationships [25]. Some of the factors underlying SRTB explored in these studies include socio-demographic factors and relationship/behavioral factors [25, 26, 29, 33, 45]. A Kenyan study conducted among university of Nairobi students describes younger age, being female, tobacco use and a previous diagnosis of a sexually transmitted infection (STI) as correlates of SRTB [33]. The University of Nairobi study is quantitative therefore, limiting understanding of the mechanisms underlying the drivers reported from the participants point of view.

Altogether, it is known from previous studies [10, 24–44, 46] that university students in sSA engage in SRTB, there is paucity of data on correlates of SRTB and their connectedness among university students at the Kenyan Coast. This study seeks to provide an in-depth qualitative understanding of SRTB among university students at the Kenyan coast using a systems thinking approach.

## Methods

### Study design

A qualitative study was conducted at Pwani university in Coastal Kenya between October 31^st^ 2019 and March 16^th^ 2020. Undergraduate students aged 18-24 years were eligible. Other stakeholders including the Dean of students, Student Counsellor, and nurses working at the students’ health unit and the university HIV voluntary counselling and testing centre (VCT) were also recruited as key informants.

### Recruitment

Recruitment of undergraduate students was through snowballing whereas the other key informants were purposively selected. Initially a meeting was held with undergraduate students to introduce the study. From this meeting, a few students volunteered to participate in the study. Recruited students recommended other students deemed to be knowledgeable on the subject matter. The process was repeated with all new interviewees until saturation was reached. To explore diversity, efforts were made to ensure students recommended for participation were spread across years of study, gender, program of study and region where they came from. Students who were opinion leaders, specifically those holding leadership positions in student organizations were given preference as they were deemed to have more insight on the subject matter. For the other key informants, the Dean of students and other staff who work closely with the students and on matters around SRTB were purposively recruited.

### Data collection

We conducted in-depth interviews (IDI) with the students and other key informants. All interviews were conducted in English. Each interview lasted about an hour and was conducted at a time convenient to each key informant. Student interviews were conducted in a private room within the VCT. For convenience, stakeholder interviews were conducted in their offices where privacy was assured. Participants were each reimbursed Kenyan shillings 350 (an equivalent to about USD 3.00) to compensate for time spent. Interviews were moderated by the main author (SC) in English and permission for notes taking and audio recording was sought *a-priori*. An interview guide was earlier developed following the World Health Organization (WHO) guidelines on school-based student health surveys [47] (S1 File). Participants’ perceptions were explored using general open-ended questions followed by additional probing where appropriate. Participants’ sociodemographic data including date of birth, gender and program of study were also collected.

### Data analysis

A distribution of the study participants by their socio-demographic characteristics was done using frequencies and percentages. Audio recordings from IDIs were transcribed. To ensure the study team could not directly associate the transcripts with individual participants, identifying information was not included in the transcripts. A thematic framework analysis approach [48], was applied as follows: Firstly, the transcripts were coded in QSR NVivo 12 (QSR International Ltd, Southport, UK). Initial coding was guided by major themes from the IDIs and socio-ecological model. New codes and themes were developed on the basis of the IDI transcripts. Secondly, excerpts were reviewed to identify common themes and variant views. Codes representing similar themes were collapsed to develop fine codes. Finally, illustrative verbatim that represent each theme were selected and presented. The interview numbers used in presenting the verbatim were not known to the study participants or any other person outside the study team. To develop the CLD, first a causal structure diagram (CSD) (S2 File) was prepared based on the data (ex-post development) [23] to identify the causal relationships between the correlates of SRTB. The information from the CSD was then used to develop the CLD using Vensim software version 9.3.4 (Ventana systems Inc 2015). Where the data did not suggest presence of a connection between two variables of interest, extra connections were added based on literature, and where necessary variables renamed, to complete the causal system. In those instances, the additional connections and variables were indicated in red.

### Ethical considerations

Prior to recruitment, a written informed consent was obtained from all potential participants. Ethical clearance was granted by Pwani University Institutional Scientific and Ethics Review Committee (ERC/PhD/003/2019) and the Kenya Medical Research Institute Scientific and Ethics Review Unit (KEMRI/SERU/CGMR-C/166/3925). Additionally, administrative approvals were granted by the National Council for Science Technology and Innovation (NACOSTI/P/19/1142) and Pwani university (Ref: PU/DVCRE/RSCH/VOL.1/37).

## Results

### Characteristics of participants

IDIs were conducted among students (n = 21) and other stakeholders (PU staff [n = 5]). Of the 26 participants, the majority was female (n = 16 [62%]). The median age was 21 years (min/max; 18 – 24) and 52 years (min/max; 32 – 58) for students and other stakeholders, respectively (Table 1).

**Table 1:**
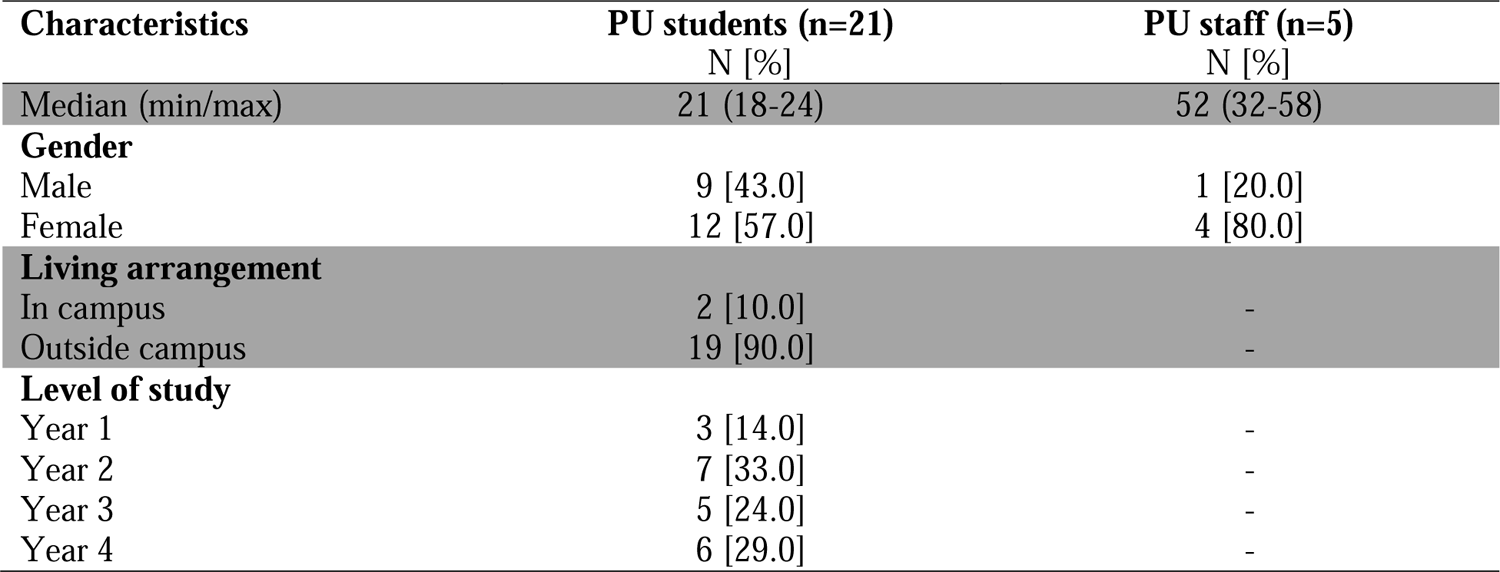
Characteristics of emerging adults at a tertiary education institution in Coastal Kenya

| Characteristics | PU students (n=21)<br>N [%] | PU staff (n=5)<br>N [%] |
| --- | --- | --- |
| Median (min/max) | 21 (18-24) | 52 (32-58) |
| **Gender** |  |  |
| Male | 9 [43.0] | 1 [20.0] |
| Female | 12 [57.0] | 4 [80.0] |
| **Living arrangement** |  |  |
| In campus | 2 [10.0] | - |
| Outside campus | 19 [90.0] | - |
| **Level of study** |  |  |
| Year 1 | 3 [14.0] | - |
| Year 2 | 7 [33.0] | - |
| Year 3 | 5 [24.0] | - |
| Year 4 | 6 [29.0] | - |

### Perceived forms of SRTB among students

Overall, participants considered unprotected sex, transactional sex, cross-generational sex, multiple sex partnerships, gender-based violence, sex under influence of alcohol/drugs, early sex debut and sharing sex toys as SRTB. Unprotected sex was considered the most common form of SRTB among students. Others were transactional sex, cross-generational sex and multiple sex partnerships (Table 2).

**Table 2:** Common forms of SRTB

| SRTB | Key informants<br>(n = 26) | Number of times<br>SRTB described |
| --- | --- | --- |
| Unprotected sex | 26 | 101 |
| Transactional sex | 25 | 136 |
| Cross generational sex | 25 | 64 |
| Multiple sex partners | 25 | 67 |
| Gender-based violence | 20 | 51 |
| Sex under influence of alcohol/drugs | 14 | 21 |
| Early sex debut | 11 | 15 |
| Sharing of sex toys | 7 | 12 |

Participants explained that most unprotected sex occurs in the context of sex under the influence of alcohol/drugs:

> “A male student… goes out with a female student. Both of them take alcohol, then they engage in the act[sex]. Of course, alcohol reduces the level of consciousness and once they have taken alcohol, they won’t even remember usage of condoms.” (Interview 1; key informant)

Participants also explained that at times students would just want to feel the pleasure of sex without the condom barrier. Where students are cohabiting, they would not see the need to use condom with their partners because they are used to each other. For students engaging in sex with older partners for financial benefits, unprotected sex would occur if the financier demanded it. At times unprotected sex was perceived to occur in the context of gender-based violence:

> “So, this male student decides to take his girlfriend for depo injection [family planning method], reason being …they are tired of using condoms, so, he wants to do it raw. So, in that scenario I would say they’re putting themselves at risk of course for one the boy is not sure if he is the only sexual partner to that lady, because they are not married … and the lady is not sure the gentleman is only having her as sexual partner.” (Interview 1; key informant)

Similarly, transactional sex was also described. Transactional sex was mostly characterized by exchange of money for sex. However, sex for grades or other favors was also described.

> “Because of financial challenges will involve themselves in sexual activities with adults of course female adults …. money for them to survive …. The same applies to ladies, they’ll get involved in… sexual activities with adults, male adults and…they may end up contracting serious diseases.” (Interview 1; key informant)

Cross-generational sex mostly occurred in the context of sugar mummy and sugar daddy relationships where sex is exchanged for money, material things or favors including good grades.

> “…particularly our lecturers, the old lecturers, it is called sex for grades, where one doesn’t attend classes, she has failed, but you find in her transcript it is written she got an A.” (Interview 10; key informant)

Multiple sexual partnerships including concurrent partners were also considered to occur in the context of transactional sex. Participants perceived that economic hardships created the need for multiple partners.

> “…Okay, our girls at Pwani university what they do they have their peer boyfriend, I mean a boyfriend of their age and then… they look for another man who will be giving them money because this boyfriend doesn’t have money. I can …call them ceremonial boyfriends…who will be financing them, herself and her boyfriend…This one mostly is an elder person, …almost their father’s age. And this is not good because these older men don’t consider using protection…” (Interview 26; key informant)

Gender-based violence was also considered common and often took the form of forced sex and predominantly involved girls as victims. A male partner would use physical force or offer alcohol to the girl with the aim of sleeping with her when she is drunk. At times, non-alcoholic drinks laced with drugs would be offered.

> “There are situations where the students engage in sex without consent from one party. So, one party is forced to accept. So, in the process they don’t even think of condoms.” (Interview 1; key informant)
>
> “They do take advantage of ladies mostly when they go to clubs. These guys have an intention of having sex with this girl… and they take her to the club and…give her some drug, or alcohol in excess … then they say “*anableki*” [becomes unconscious], so, after that you find yourself in a room with different guys so they end up having sex with you, you end up getting disease, maybe HIV or STIs.” (Interview 16; key informant)

Sex under the influence of alcohol/drugs was considered risky as it was frequently unprotected.

> “When people are into drugs they forget themselves and find themselves doing that act [sex] may be not protected then they can contract [STIs].” (Interview 17; key informant)

It was perceived that those engaging in SRTB are likely to have started sexual activities at an early age. Some participants felt that sexual debut could occur even among nine-year olds.

> “The age at which for lady to begin early sex even can start at the age of 9 years and for the men maybe 12 years.” (Interview 5; key informant)

Sharing of sex toys among lesbians was also reported and could facilitate transmission of STI including HIV.

> “The lesbians…use those tools [toys]. …maybe it is a group of lesbians and maybe they are using that one vibrator and you never know if one has the STI through that they may contract.” (Interview 12; key informant)

### Risk and protective factors for SRTB among students

The respondents identified a number of risk and protective factors for SRTB. Based on the ecological model, most of the reported risk factors operated at the individual level followed by those at social family and peer level. Similarly, most of the reported protective influences operated at the individual level (Fig 1 and Table 3).

**Fig 1:**
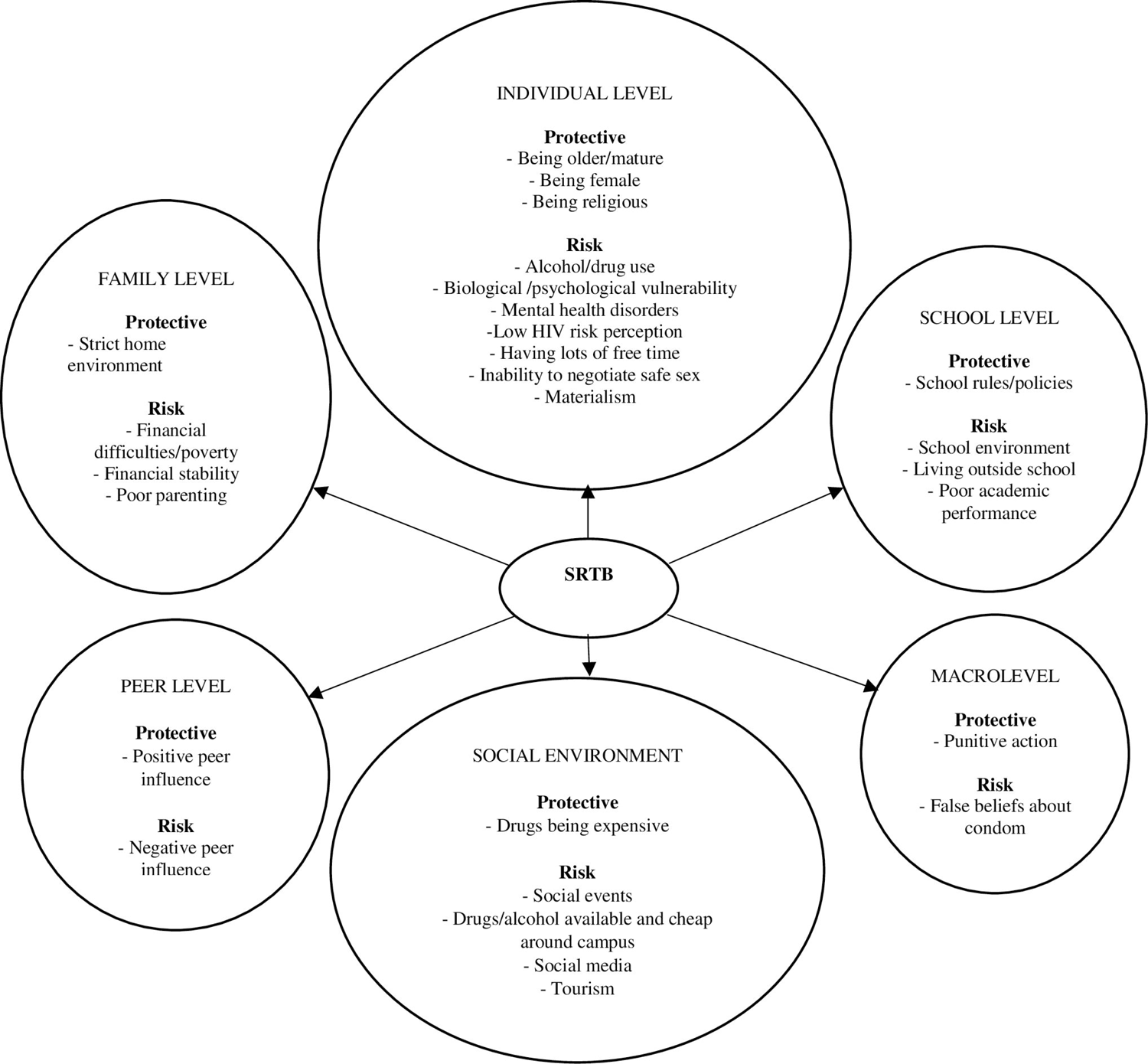
A flow diagram of emerging adults’ and other stakeholders’ perceived risk and protective factors for sexual risk taking behaviour among emerging adults at a tertiary education institution in Coastal Kenya

**Table 3:** Risk and protective factors of sexual risk-taking behaviour and exemplifying statements

| Factor level |  | Factor | Exemplifying statement |
| --- | --- | --- | --- |
| Individual | Risk | Alcohol/drug use | ...People are normally raped because they... served juice which maybe has been laced with alcohol or whatever, in the morning they come crying .....for PEP [Post exposure prophylaxis] ...rape has taken place and ... |
|  |  | Biological vulnerability | ...some do it[sex] for pleasure, some people who drive pleasure by having many boyfriends... they want the sex they don't want the commitment...that is why disease and pregnancy [are common] ... |
|  |  | Mental health disorders | ...when a person is depressed they may not be in a position to ... make the right decision ... and they may engage more in risky behaviour, they might have reached a point where they don't care about anything and therefore taking measures to have safe sex may not be in their minds in that depressed state... |
|  |  | Low HIV risk perception | ...some don't simply because they have a girlfriend and that girlfriend... may not want. For example, I live with a girl in my house, you know condom...becomes boring because you are with her there every now and then... but to others they may stop using simply a lady can tell you why have you decided to use a condom on me don't you trust me... |
|  |  | Having lots of free time | ...Idleness ... because there is nothing you do so some will ...go hung around...they date or sleep somewhere with somebody she or he doesn't know their [HIV] status...and the end of it all you find that the person you were with may be had HIV... |
|  |  | Inability to negotiate safe sex | ... yes, they will do it [unprotected sex] willingly...what this boy will tell you is 'if you don't do it then I will leave you' then others fear they don't have that power to refuse. |
|  |  | Materialism | ... this occurs mostly to the girls, like if your parents are not giving you enough money to support you in the university or you are greedy you see may be the money they are giving you is for food, just for food but you want more, you want to do your hair, you want your house to look good, to have good furniture, you want to go for vacation every weekend like you want to live a good life and you don't have the money for that good life and your parents don't have the money, so you find that they will go look for those sponsors because those sponsors like the old men usually want the young girls and they will do anything for you as long as you are ... having sex with them... they will give you any kind of money you want... |
|  | Protective | Being older/mature | ...the older students some of them are mature... most of them know what is good and what is bad for them but the younger students they have no limit they are not guided to know what is good and what is bad they take everything for fun |
|  |  | Being female | ...it is not easy for a girl because ...they do the sexual [activities] from the heart but the men do it for pleasure just to satisfy their desire that time and they go ...so it is not easy for a girl to engage in sex easily but for the male students it is very easy ... |
|  |  | Being religious | ...A religion such as Muslim...you'll find their religion has limited the girls to engage in sexual activities before marriage... |
| Social environment | Risk | Social events | ...This student decided to go for freshers' night [party for new students]. And I had warned her earlier, I had told her that thing isn't beneficial to you in any case, but she forced out and attended. So, when ...she went home she called me and told me I'm feeling unwell. I asked why? How are you feeling? I'm feeling like vomiting, I have stomach ache that is crampy. Since when? Around 3 days ago. Then I told her those looks like signs of pregnancy so please go to the hospital, get a kit and test. So, when she did that the results were positive... |
|  |  | Drugs available & cheap in campus | ...They are several, they are just around the place, they can get them very easily, and they are being sold very cheaply... |
|  |  | Social media | ... [they find the sponsor/sugar daddy] on face book... |
|  |  | Tourism | You know this is coast there are rich people[tourists] around the beaches... some people do not have the boundaries of knowing you are dating a person almost your father age... an old guy approaches you he gives you his number "anaanza kukukatia unaingia box" [he seduces you and you give in] ...some people can go for it because of the money |
|  | Protective | Drugs being expensive | Some inject but ...they rarely do it... you know...injection drugs are very expensive... so most of them normally students don't afford it... |
| Peer | Risk | Negative peer influence | Girls especially, they are not able to make quite good decisions, and this led them to being harassed or lured into sex and latter you'll find out they have not used protection |
|  | Protective | Positive peer influence | ...you are talking about the same issues you can share your personal issues or you can share to the group what you went through and how you have become, it helps |
| Family | Risk | Financial difficulties | Because of financial challenges will involve themselves in sexual activities with ... female adults to get money ... to survive.... The same applies to... university ladies, they'll get involved in... sexual activities...with male adults and.... end up contracting serious diseases |
|  |  | Financial stability | Then there is HELB money [student loan] that comes in ...comrades [students] gather then they so many parties.... like parting and in those parties, alcohols are the accompaniment, sometimes in the rentals we hear them shouting... but eventually I think illicit sex takes place... |
|  |  | Lack of parental monitoring | That one [SRTB] will mostly depend with parental care you have at home... at an early age below 18 it is mostly influenced by the lack of proper parenting... |
|  | Protective | Strict home environment | When you are at home the chances of being lured [into SRTB] are very low...when your home ground is so strict... |
| School | *Risk* | School environment | Ladies end up having sex with different guys because ... she loved this guy who broke her heart, so she ends up sleeping with anyone... |
|  |  | Living outside school | If they are out there [residence outside campus] .... they have the freedom... then it means they can involve in anything and as youth we know there are many challenges particularly on alcohol and drug abuse, matters of sexual behaviour... |
|  |  | Poor academic performance | ...they make...poor judgment in their academics this results to their failure and causes them to engage in sexual behaviour which are risky... |
|  | *Protective* | School policies/rules | ...the university issue condoms to all students free... |
| Macro | *Risk* | False belief about condom | There is that notion that when you put on a condom, the sexual effect is not as sweet as when you are not using... |
|  | *Protective* | Punitive action | In campus you will face the consequences definitely you would like to engage [in SRTB] out there... |

The CLD demonstrated the interconnectedness of the correlates of SRTB as illustrated by the numerous feedback mechanisms that influence SRTB. Transactional sex, cross generational sex and multiple sex partnerships formed a reinforcing feedback loop in which each had an incremental effect on the other. The loop was driven by sex tourism, visibility of sugar mummies/daddies on social media, financial difficulties, poor academic performance, acceptance of sex drive in women and men, traditional gender norms, liberal sexual norms and the desire to acquire material things. However, being mature reduced transactional sex, which consequently had the same effect on all components of the loop.

Similarly, societal acceptance of unsafe sexual practices seemed to increase unprotected sex and formed a reinforcing loop with it. Transactional sex, having lots of free time, sexual violence, sex under influence of alcohol/drugs and false beliefs about condoms increased unprotected sex therefore introducing an incremental effect on the reinforcing loop between unprotected sex and societal acceptance of unsafe sex. However, adherence to religious principles, parental control and ability to negotiate safe sex reduced unprotected sex. (Fig 2).

**Fig 2:**
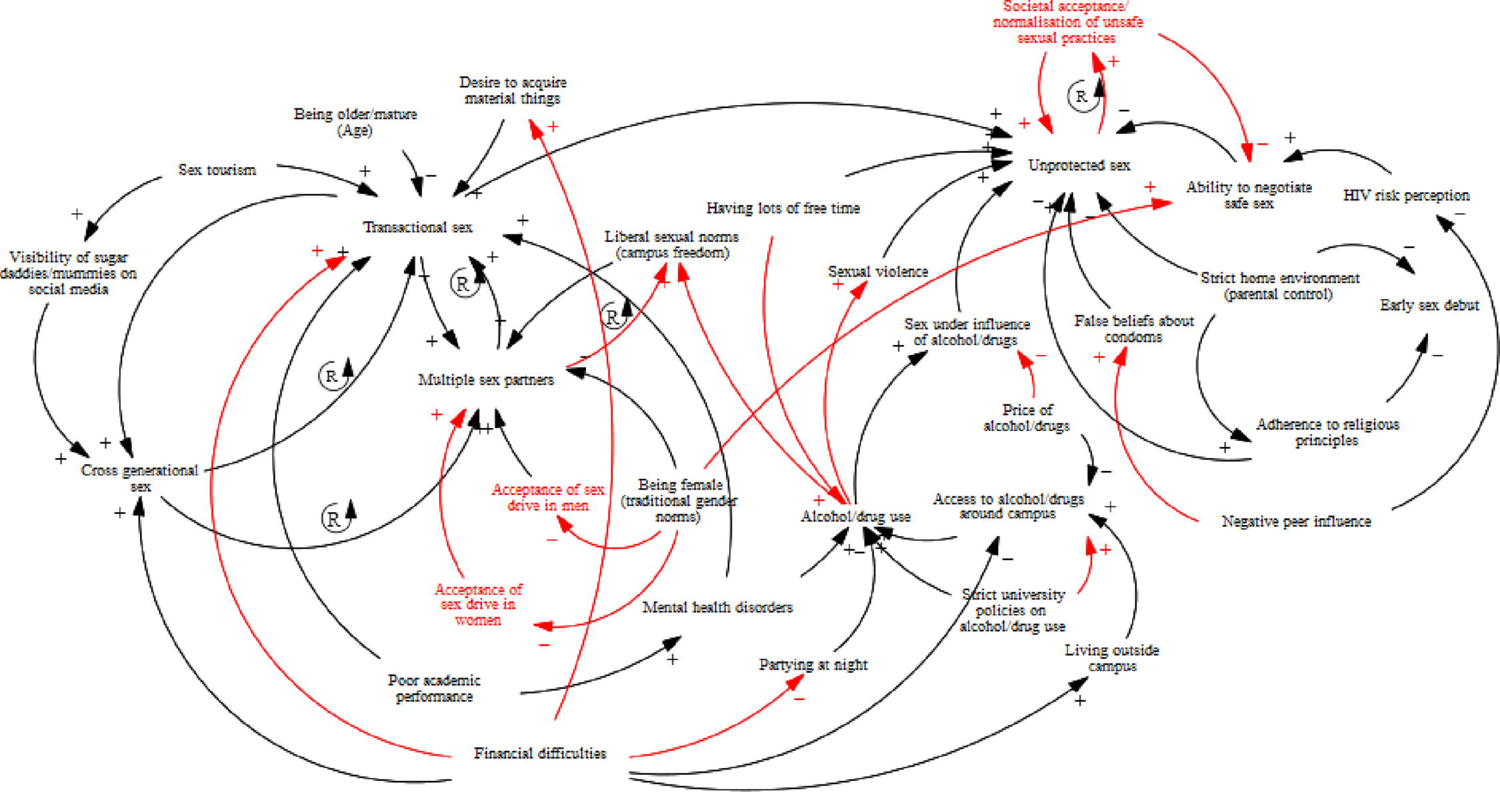
A causal loop diagram showing risk and protective factors that impact sexual risk-taking behaviour among emerging adults at a tertiary education institution in Coastal Kenya. Arrows and variables in red have been added based on literature to make the causal loop diagram complete

### Individual level factors

#### Risk factors

##### Alcohol/drug use

Participants explained that alcohol/drug use was a contributing factor to SRTB. In some instances, male students would buy alcohol for girls with the intention of engaging in sex with them once they get drunk. In these circumstances the male partner is taking advantage of the drunk state of the girl. Where the girl is still conscious enough to resist, the male partner would use force. Participants also narrated that at times the intention to take advantage of one partner was not there but sexual risk behaviour would still happen because alcohol impedes their decision making. In addition, it was revealed that risky sexual behaviors were happening after abusing drugs other than alcohol.

> “People are normally raped because they… served juice which maybe has been laced with alcohol or whatever, in the morning they come crying for PEP [Post exposure prophylaxis] …rape has taken place and ….” (Interview 25; key informant).
>
> “They have group sex, and they have it when they are also drunk, so I think when somebody is drunk … they cannot decide very well, even if they wanted to use a condom for example, they will not use it. So, and the gay also some of them don’t even consider the condom they just do it without condom. And then they come back and tell me they want to access the PEP…” (Interview 26; key informant).

##### Biological/psychological vulnerability

Participants felt that biological factors including sex and personal characteristics like being inclined to have fun, curiosity/wanting to explore and wanting to be famous were contributing to SRTB by emerging adults. Interestingly, both male and female sex were reported to increase vulnerability to SRTB though most participants were of the view that being male was a contributing factor to SRTB.

> “…some do it[sex] for pleasure, some people who drive pleasure by having many boyfriends… they want the sex they don’t want the commitment…that is why disease and pregnancy [are common].” (Interview 14; key informant).

##### Mental health disorders

Mental health disorders especially depression and anxiety were perceived to be pushing EmA to engage in alcohol and subsequently SRTB.

> “…when a person is depressed they may not be in a position to … make the right decision … and they may engage more in risky behaviour, they might have reached a point where they don’t care about anything and therefore taking measures to have safe sex may not be in their minds in that depressed state.” (Interview 23; key informant).

##### Low HIV risk perception

Participants reported that EmA who believe have low chances of contracting HIV were more likely to engage in SRTB. Participants reported that EmAs would engage in sex without condom because they do not like using condom, have negative perception about it or they are used to their partner so do not see the need to use condom. Further, low HIV risk perception among EmA was exemplified by other risky activities/beliefs that they were reported to engage in including deep kissing and fear of pregnancy but not HIV infection.

> “…some don’t simply because they have a girlfriend and that girlfriend… may not want. For example, I live with a girl in my house, you know condom…becomes boring because you are with her there every now and then… but to others they may stop using simply a lady can tell you why have you decided to use a condom on me don’t you trust me?” (Interview 20; key informant).

##### Having a lot of free time and inability to negotiate safe sex

These were also seen to contribute to SRTB. Participants explained that students with a lot of free time tend to spend that time thinking about and engaging in SRTB.

> “Idleness … because there is nothing you do so some will …go hung around…they date or sleep somewhere with somebody she or he doesn’t know their [HIV] status…and the end of it all you find that the person you were with may be had HIV.” (Interview 5; key informant).
>
> “Yes, they will do it [unprotected sex] willingly…what this boy will tell you is ‘if you don’t do it then I will leave you’ then others fear they don’t have that power to refuse.” (Interview 12; key informant).

##### Materialism

Participants revealed that the love for money and material things was pushing especially ladies into transactional sex just to maintain their desired lifestyle.

> “… this occurs mostly to the girls, like if your parents are not giving you enough money to support you in the university or you are greedy you see may be the money they are giving you is for food, just for food but you want more, you want to do your hair, you want your house to look good, to have good furniture, you want to go for vacation every weekend like you want to live a good life and you don’t have the money for that good life and your parents don’t have the money, so you find that they will go look for those sponsors because those sponsors like the old men usually want the young girls and they will do anything for you as long as you are … having sex with them… they will give you any kind of money you want.” (Interview 12; key informant).

### Protective factors

#### Being older/mature

Participants explained that unlike the younger/junior students, mature students are focused on future life and academics and generally know what is good for them so they are more likely to make right decisions regarding sexual matters.

> “The older students some of them are mature… most of them know what is good and what is bad for them but the younger students they have no limit they are not guided to know what is good and what is bad they take everything for fun.” (Interview 4; key informant).

#### Being female

It was revealed that males are more prone to careless sex because they do it for pleasure with no emotional attachment unlike girls.

> “It is not easy for a girl because …they do the sexual [activities] from the heart but the men do it for pleasure just to satisfy their desire that time and they go …so it is not easy for a girl to engage in sex easily but for the male students it is very easy.” (Interview 12; key informant).

#### Being religious

Religion was perceived to help reduce SRTB since most religions prohibit sex before marriage.

> “A religion such as Muslim…you’ll find their religion has limited the girls to engage in sexual activities before marriage.” (Interview 10; key informant).

### Social environment factors

#### Risk factors

##### Social events

Participants narrated that social events organized both inside and outside campus provided an opportunity for students to engage in SRTB. Freedom enjoyed by the students especially outside campus was perceived to facilitate SRTB during the social events.

> “This student decided to go for freshers’ night [party for new students]. And I had warned her earlier, I had told her that thing isn’t beneficial to you in any case, but she forced out and attended. So, when …she went home she called me and told me I’m feeling unwell. I asked why? How are you feeling? I’m feeling like vomiting, I have stomach ache that is crampy. Since when? Around 3 days ago. Then I told her those looks like signs of pregnancy so please go to the hospital, get a kit and test. So, when she did that the results were positive.” (Interview 1; key informant).

##### Drugs/alcohol available and cheap around campus

Participants felt that alcohol and other drugs were readily available and cheap around campus which increased the risk of engaging in sex while under influence of the drugs.

> “They are several, they are just around the place, they can get them very easily, and they are being sold very cheaply.” (Interview 26; key informant).

##### Social media

Social media use was seen to predispose students to SRTB. Participants narrated how students connect with their sugar mummies/daddies on Facebook. Further, they explained that students watch pornography which fuels their curiosity to try it out.

> “[they find the sponsor/sugar daddy] on face book.” (Interview 12; key informant).

##### Tourism

It was perceived that tourism contributed to SRTB. Since the university is located along the Kenyan Coast, some tourists, usually old, along the beaches were targeting female students. The students would in turn give in as they expect to benefit financially.

“You know this is coast there are rich people[tourists] around the beaches… some people do not have the boundaries of knowing you are dating a person almost your father age… an old guy approaches you he gives you his number “*anaanza kukukatia unaingia box*” [he seduces you and you give in] …some people can go for it because of the money.” (Interview 14; key informant).

### Protective factors

#### Drugs being expensive

While participants felt that drugs were easily accessible and cheap, especially palm wine, locally known as “mnazi”, there were also expensive drugs that students could not access. This keeps them sober hence reducing the chances of engaging in SRTB.

> “Some inject but …they rarely do it… you know…injection drugs are very expensive… so most of them normally students don’t afford it…” (Interview 10; key informant).

### Peer level factors

#### Risk factors

##### Negative peer influence

Most participants felt that close friends can influence others into risky sexual encounters. Some of the negative influences include non-condom use during a sexual encounter while drunk.

> “Girls especially, they are not able to make quite good decisions, and this led them to being harassed or lured into sex and latter you’ll find out they have not used protection.” (Interview 10; key informant).

### Protective factors

#### Positive peer influence

Sharing experiences by peers was seen to help them overcome common challenges.

> “… you are talking about the same issues you can share your personal issues or you can share to the group what you went through and how you have become, it helps.” (Interview 14; key informant).

### Family level factors

#### Risk factors

##### Financial difficulties/poverty

Participants narrated that due to financial challenges, students would engage in SRTB in order to cater for their needs.

> “Because of financial challenges will involve themselves in sexual activities with … female adults to get money … to survive…. The same applies to… university ladies, they’ll get involved in… sexual activities…with male adults and…. end up contracting serious diseases.” (Interview 1; key informant).

##### Financial stability

Though financial difficulties were perceived to predispose to SRTB, financial stability was equally perceived to have the same effect. Financially stable students or those with well-off friends were more predisposed to SRTB because they could afford the expenses associated with such activities including going out for social events with their partners.

> “Then there is HELB money [student loan] that comes in …comrades [students] gather then they so many parties…. like parting and in those parties, alcohols are the accompaniment, sometimes in the rentals we hear them shouting… but eventually I think illicit sex takes place.” (Interview 25; key informant).

##### Lack of parental monitoring

Decision making on sexual matters was seen to be influenced by parental care that one receives at an early age. Lack of close parental monitoring was seen to negatively influence decision making regarding sexuality leading to SRTB.

> “That one [SRTB] will mostly depend with parental care you have at home… at an early age below 18 it is mostly influenced by the lack of proper parenting.” (Interview 2; key informant).

### Protective factors

#### Strict home environment

A strict home environment where there is close parental monitoring was perceived to offer protection from being lured into SRTB.

> “When you are at home the chances of being lured [into SRTB] are very low…when your home ground is so strict.” (Interview 24; key informant).

### School level factors

#### Risk factors

##### School environment

Participants explained that some factors within the school environment could predispose students to SRTB. Most participants pointed out that sexual relationships in campus are temporary and sometimes students find themselves engaging in SRTB to cope with heart break following break-up of a relationship. Similarly, campus freedom that allows students to have sexual relationships and weak security systems at campus were blamed for SRTB among students.

> “Ladies end up having sex with different guys because … she loved this guy who broke her heart, so she ends up sleeping with anyone.” (Interview 16; key informant).

##### Living outside school

Compared to residence within campus, participants were of the view that residing outside campus exposed students to SRTB. This is because the campus freedoms are there but are practiced within the rules in campus. This is not the case outside campus where the campus rules do not apply.

> “If they are out there [residence outside campus] …. they have the freedom… then it means they can involve in anything and as youth we know there are many challenges particularly on alcohol and drug abuse, matters of sexual behaviour.” (Interview 22; key informant).

##### Poor academic performance

Participants explained that when students perform poorly in academics, some get depressed and end up engaging in SRTB. Female students performing poorly would be ready to have risky sexual relations with lecturers to rescue themselves.

> “… they make…poor judgment in their academics this results to their failure and causes them to engage in sexual behaviour which are risky.” (Interview 10; key informant).

### Protective factors

#### School rules or policies

Participants felt that school policies and rules which directly affects those living in campus stops students from engaging in SRTB. Examples of policies include restriction of alcohol use and provision of mental health counselling services on campus which keeps frustrated students from engaging in SRTB as a way of coping. Similarly, provision of condom on campus was mentioned.

> “… the university issue condoms to all students free.” (Interview 25; key informant).

### Macro level factors

#### Risk factors

##### False beliefs about condoms

Participants explained that some students were not using condom because of certain beliefs. Some believed that condoms reduce sexual pleasure while others believe condoms will burst.

> “There is that notion that when you put on a condom, the sexual effect is not as sweet as when you are not using…” (Interview 24; key informant).

### Protective factors

#### Punitive action

It was revealed that negative consequences imposed on students for engaging in SRTB makes them shy away from it. Participants explained that students caught engaging in sex in public spaces within campus due to influence of alcohol face consequences including expulsion from the university.

> “In campus you will face the consequences definitely you would like to engage [in SRTB] out there.” (Interview 23; key informant).

## Discussion

In this study we set out to understand the perceived SRTBs among EmA and associated risk factors. Our findings show that EmA and other stakeholders perceived SRTB to be common at an institution of higher learning in Coastal Kenya, especially unprotected sex, transactional sex, cross generational sex, multiple sexual partnerships, gender-based violence, sex under influence of alcohol/drugs, early sex debut, and sharing sex toys. Biological/psychological vulnerability, negative peer influence, alcohol/drug use and financial difficulties were most frequently described as drivers of SRTB in this population.

Unprotected sex, sex under the influence of alcohol/drugs and gender-based violence were described to be common among the EmA. Further, the CLD shows that the three are interlinked with gender-based violence and sex under influence of alcohol/drugs increasing unprotected sex. Consistent with our findings, previous studies from sSA have found unprotected sex to be the most common SRTB which underscores its importance [14]. Indeed, a recent systematic review and meta-analysis of EmA age 18-25 years from Africa found that non-condom use had the highest pooled prevalence estimate of 46% [14]. Participants explained that alcohol/drugs impair judgement hence when engaging in sex while under influence, EmA may not bother using a condom and if they do, it will not be used correctly. In addition, gender-based violence seems to be fueled by alcohol/drug use in this population. Social events organized both on and off campus provide an opportunity for alcohol/drug use. In these circumstances, perpetrators of gender-based violence would offer alcohol/drugs to the targeted victims then assault them sexually. It is possible that alcohol/drug use and the social events could be behind unprotected sex, sex under influence of alcohol and gender-based violence in this population as it has been found in other studies [14] and confirmed in the CLD where alcohol/drug use increases sex under influence of alcohol/drugs which in turn increases unprotected sex. There is need for universities to strengthen mental health support services including counselling to help reduce alcohol/drug use among EmA [49].

Transactional sex, cross generational sex and multiple sex partners were perceived to be common amongst EmA and mainly occurred in the context of sex in exchange of money. Further, the CLD shows that the three SRTBs are interlinked in a reinforcing loop in which transactional sex increases cross generational sex which then increases multiple sex partnerships that further increases transactional sex in a vicious cycle. A recent review equally found transactional sex, cross-generational sex and multiple sex partnerships to be prevalent among EmA in sSA [14]. Similarly, financial difficulties came out strongly among participants as a risk factor for transactional sex. The Kenyan government rolled out free primary education in 2003 which substantially increased primary school enrollment [50]. Additionally, the government has in recent years been implementing the 100% transition policy which aims to ensure all primary school pupils progress to secondary school and later either join university or middle level colleges [51]. These two factors may have contributed to the steady increase in public university enrollment from 442,000 in 2015 to 1,000,000 in 2018 [52]. Interestingly, government funding in the higher education sector has either stagnated or reduced [52]. Though tuition fees in public universities remain affordable, public universities have had to find ways to bridge the funding gap. Many public universities have passed certain costs to parents/students, example is industrial attachment which students are currently paying for directly. Apart from universities passing costs to students, students’ financial burden has further been increased by the fact that the higher education loans board, a government body that provides loans to university students to help them fund their education, has not been able to cope with the increasing number of students [53]. Further, loan recovery has been a challenge due to the high youth unemployment rate in Kenya [53]. Disbursements have therefore been inadequate for those who secure the loan while many miss out altogether [53]. Students welfare has also posed a challenge as most students are accommodated outside campus therefore paying rent at market price. It is possible that the fore mentioned factors may have rendered university education expensive hence many parents struggle to support their children. Under these circumstances, it is plausible that some students engage in transactional sex, cross generational sex and multiple sex partnerships to cater for their needs at the university. However, materialism and peer pressure were also described to be risk factors for SRTB. It is possible that the desire to acquire material things to maintain an expensive life style like their financially-stable peers was pushing EmA to engage in transactional sex, cross generational sex and multiple sex partnerships like it has been demonstrated in previous studies [54]. The CLD confirms that financial difficulties and desire to acquire material things directly increase transactional sex, cross generational sex and multiple sex partnerships. Kenyan universities need to enhance sexual education programs for students so that they can be empowered and avoid the urge to engage in SRTB for financial gain [55]. Although transactional sex mainly involved exchanging sex for money, sex for favorable grades was also reported. A previous study among adolescents in this setting equally revealed that transactional sex was common but it did not involve exchange of sex for favorable grades despite the fact that some of the adolescents were school-going [54]. It seems exchange of sex for favorable grades is a common form of transactional sex among young people in universities but not those in high schools or primary schools. Kenyan universities have administrative procedures to deal with sex for grades. It is probably time for the universities to review those procedures and strengthen them to ensure such cases are reduced.

One strength of this study is the use of CLD to map the SRTB system in this population. An important limitation of this study is that it only involved participants from one university. It remains unclear if their views could differ from those in other universities given the geographical differences.

## Conclusion

This study explored SRTB among EmA at a public university in Coastal Kenya. Consistent with findings from studies among EmA in general population, our study shows that EmA are engaging in SRTB with unprotected sex perceived to be the most common and seems to be driven by individual level factors especially alcohol/drug use. In a unique way, our study utilizes systems thinking approach to build on existing literature by highlighting the perceived drivers of SRTB among university students and their interlinkages. Enhancing sexual education programs for university students in Kenya and strengthening support systems including counselling for those using alcohol/drugs may help reduce SRTB among EmA in universities in Kenya.

## Data Availability

The data underlying the results presented in the study are readily available upon request from the corresponding author upon request because it contains potentially identifying information

## Acknowledgement

The authors would like to thank all participants for volunteering to take part in this study. Further, the authors are grateful to Karabu Ngombo (CGMR-Coast, Kenya) and Judith Tumaini Dzombo (CGMR-Coast, Kenya) for their help during data collection. This manuscript was submitted for publication with permission from the Director of the Kenya Medical Research Institute/Wellcome Trust Research Programme.

## Supporting information

S1 File. Interview guide

S2 File. Causal structure diagram

## Author contributions

Conceptualization - SC, AH, AAA (Amina Abubakar), KM, SA, AA (Alice Anika), FI, VK, OA Data curation – SC, AAA, VK, KM, EO, IM, CN

Formal analysis – SC, AAA, KM Funding acquisition – AH, AAA Investigation – SC, KM, SA, AH

Methodology – SC, AH, KM, SA, AAA Project administration – OA, AAA, AH Resources – SA, KM, AAA, AA, FI Supervision – KM, SA, AH, OA

Validation – KM, SA, AH, AA, VK, OA, FI, AAA

Visualization – SC, VK, EO, IM, CN Writing-original draft preparation – SC

Writing – review and editing – KM, SA, AH, AAA, VK, OA, AA, FI, EO, IM, CN

