## Supplementary file 1 for "Sexual risk-taking behavior amongst emerging adults in a tertiary institution of learning in Coastal Kenya: A qualitative study of stakeholders’ perspectives using causal loop mapping"

**Relationship between neurocognition, mental health disorders and sexual risk- taking behavior with HIV infection among emerging adults in a tertiary institution in Coastal Kenya**

### S1 File: Guide for in-depth interviews with key informants

### Instructions to interviewer

- Greet the participants
- Introduce yourself
- Give background information about the study. Carefully read through the informed consent form and answer any questions. Give information about the interview.
- Assure the participants that he/she does not have to participate if he/she does not want to.
- Ask for approval to participate in the interview.
- Tell the participants that he/she can stop the interview any time they wish.
- Assure confidentiality.
- Make sure the participants know what the tape recording procedure is. Ask for approval.
- Ask if the participant has any questions before the interview.
- If the participant agrees to take part please ensure they sign the consent form.
- If the participant agrees to take part please ensure they sign two copies of the consent form, give them a copy and keep one copy for our records.
- Check the tape recording equipment.
- Start the interview.

At the start of the interview, do not record respondent’s name or any identifier; just indicate the date, time, gender, age and educational level of the person being interviewed. Lastly record where the interview was taking place.

1. In your opinion what do you think are some of the challenges that young people/emerging adults/ students in this university face?

Probe on the following if not mentioned

a) In terms of mental health, what would you say are the challenges/problems experienced by emerging adults/undergraduate students in this university? (Ask for a description with specific examples)
b) In terms of sexuality, what would you say are the challenges/problems experienced by emerging adults/students in this university? (Ask for a description with specific examples)
c) In terms of alcohol consumption, what would you say are the challenges/problems experienced by emerging adults/students in this university?
d) In terms of drug use, what would you say are the challenges/problems experienced by emerging adults/PU students in this university?

Sometimes emerging adults/students engage in behaviour or habits likely to put them at risk of contracting STIs (including HIV) and unwanted pregnancies. In your opinion, which are the examples of such behaviours that emerging adults/students in Pwani university engage in?

(For each SRTB probe accordingly so that the discussion follows naturally)

I would like us now to discuss in more detail. Drawing from your experiences and opinions talk about the following habits/behaviours in detail with a focus on young people here in the university.

Alcohol use behaviour

Alcohol use is one of the forms of risky behaviour. Here in PU which are the forms of alcohol use behaviours that you think pose a risk of contracting STIs?

Probes:

- Mention the local names of the types of alcohol consumed by emerging adults in PU
- Can you share some examples (but do not mention names of persons involved?)
- Who in particular is likely to engage in these forms of behavior (in terms of gender/age or other socio-demographic characteristics)?
- Why, in your opinion, are they engaging in this behaviour

Sexual Behavior

We hear that at times some students in PU engage in certain forms of sexual behavior that are risky for their health. In your opinion, which are some of these sexual activities taken by students in PU?

Probes:

- Can you share some examples (but do not mention names of persons involved)?
- Who is particularly like to do this (in terms of gender/age or other socio-demographic characteristics)?
- Why, in your opinion, are they engaging in this behaviour

*Notes: (Moderator think of: early sexual debut, multiple sexual partnerships, cross generation (sugar daddy/mummy) or transactional sex, condom non-use, if it’s possible to probe for various forms of contraceptive methods)*

Smoking Behavior

Smoking is another type of risky behavior that young people engage in.

*Think about all forms of smoking behavior and describe which of these students in PU may engage in?*

Probes:

- Mention the local names of what is commonly smoked.
- Who is particularly likely to do this (in terms of gender/age or other socio-demographic characteristics)?
- Why, in your opinion, are they engaging in this behaviour

*Notes: (Moderator think of: age of onset, local names/slang cigarette use, can they identify brand names, forms of smoking such as Sisha, pipes etc)*

Drug Use Behavior

Drug use is one of the risk behaviors reported among young people in Kenya and also here in PU. *What do you think are some of these drugs used here in PU by students? Mention the local names of these drugs*

Probes:

- How are these drugs used? Can you share some examples (but do not mention names of persons involved)?
- Who in particular is likely to engage in these forms of behavior (in terms of gender/age or other socio-demographic characteristics)?
- Why, in your opinion, are they engaging in this behaviour?

*Notes: (Moderator think of: local names, routes like injecting sniffing smoking chewing), where they are accessed)*
