## Supplementary file 2 for "Sexual risk-taking behavior amongst emerging adults in a tertiary institution of learning in Coastal Kenya: A qualitative study of stakeholders’ perspectives using causal loop mapping"

S2 File: Casual structure diagram

Being religious (Adherence to religious principles)

⁺	‒

Sex under influence of alcohol/drugs

Unprotected sex

Unprotected sex

Transactional sex

Unprotected sex

Tourism (Sex tourism)

Cross generational/transactional sex

Cross generational/transactional sex

Unprotected sex

Unprotected sex

Sex under influence of alcohol/drugs

Sex under influence of alcohol/drugs

Early sex debut

Unprotected sex

Sex under influence of alcohol/drugs

SRTB

SRTB

Multiple sex partners

Sex under influence of alcohol/drugs

Unprotected sex

Unprotected sex

Alcohol/drug use

Sex under influence of alcohol/drugs

Punitive action

False belief about condom

Living outside campus

School environment

Lack of proper parenting

Financial stability

Positive peer influence

Negative peer influence

Inability to negotiate safe sex

Unprotected sex

Having lots of free time

Unprotected sex

Low HIV risk perception

Alcohol/drug use

Mental health disorders

Unprotected sex

Being female (Gender norms)

Multiple sex partners

Unprotected sex

Multiple sex partners

Transactional sex

Transactional sex

Alcohol/drug use

Being older/mature (Age)

Poor academic performance

Mental health disorders

Financial difficulties

Unprotected sex

Unprotected sex

Transactional sex

Transactional sex/cross generational sex

Cross generational sex

Forced sex/gender violence

Transactional sex

Social events (Partying at night)

⁺	⁺

Drugs expensive (Price of alcohol/drugs)

⁺	‒

⁺	⁺

Drugs available & cheap around campus (Access to alcohol/drugs)

Transactional sex

Multiple sex partners

⁺

Social media (Visibility of sugar mummies/daddies on social media)

⁺

⁺	⁺

⁺

⁺	⁺

⁺

‒

‒

⁺

⁺	⁺

Biological vulnerability (sexual drive)

‒	⁺

Strict home environment (Parental control)

⁺	⁺	‒

⁺	⁺

⁺

⁺

‒

School rules/policies (strict university policies on alcohol/drug use)

⁺

⁺

Materialism (Desire to acquire material things)

⁺

‒	‒
